# Occupational falls by site of occurrence within the workplace in Japan

**DOI:** 10.1101/2025.04.27.25326535

**Authors:** Kazuhiko Watanabe, Sora Hirohashi, Tomohiro Yoshimi, Masayoshi Zaitsu

## Abstract

**Background:** Little is known about the distribution of specific sites where occupational falls frequently occur within the workplace. This study aimed to examine the distribution of occupational falls by sites of occurrence in Japan.

**Methods:** National data of occupational falls resulting in absences of four or more days in 2023 were extracted from a website managed by the Ministry of Health, Labour and Welfare, Japan. Fall sites were classified as either outdoor or indoor. Indoor sites were further categorized as walkways, work platforms/walking planks, indoor stairs, or other sites. Walkways and work platforms/walking planks were defined as indoor level surfaces. The distribution of occupational fall by sites was described.

**Results:** Among all occupational falls, 63.2% occurred at indoor sites (22,780/36,058). The most frequently reported site was indoor level surfaces, accounting for 49.0%, while indoor stairs accounted for 5.9%. When stratified by sex, 65.9% occurred at indoor sites among female workers while 59.0% occurred among male workers. The difference was largely attributable to a higher proportion of falls on indoor level surfaces among females (52.7% in females and 43.5% in males).

**Conclusions:** In Japan, a substantial proportion of occupational falls occurred at indoor sites, particularly on indoor level surfaces.

## Introduction

The global burden of work-related health problems is substantial, accounting for approximately 180 million disability-adjusted life years lost each year and resulting in significant economic losses.^1^ Globally, approximately 2.9 million work-related deaths occur annually, with an estimated 2.6 million due to occupational diseases and 320,000 due to occupational injuries.^1^ This persistent burden highlights the urgent need for more robust measures to safeguard health and safety in the workplace.^2^

Reducing occupational injuries remains a critical public health priority. In Japan, efforts to improve workplace environments have been consistently emphasized. Meanwhile, the growing proportion of older workers has raised concerns about a potential increase in occupational injury risks.^3^ In particular, occupational falls and subsequent fall-related injuries are the most common type of workplace accident resulting in absences of four or more days. In 2023, fall-related injuries required an average of 48.5 days of leave.^4^

In addition to health consequences, such absences reduce the available workforce. Moreover, in Japan, approximately 80% of wages are compensated by worker’s accident compensation insurance starting from the fourth day of leave, underscoring the economic importance of preventing occupational falls.

However, little is known about the specific locations within the workplace where such incidents most frequently occur. Recent studies have indicated the role of intrinsic rather than environmental factors in occupational falls, including static balance function and lifestyle-related factors.^5,6^ If occupational falls are found to occur most frequently in controlled environments—such as indoor level surfaces—this would emphasize the need to shift the focus beyond environmental modifications and toward addressing intrinsic factors, such as physical functioning.

This study aimed to clarify the distribution of occupational falls by site of occurrence. We examined the proportion of falls occurring on indoor level surfaces using national data on occupational accidents and diseases in Japan.

## Materials and Methods

### Data source

Data were obtained from the national registry of the Reports of Occupational Accidents and Diseases, also known as the *Reports of Worker Casualties*. Workers who experienced occupational falls resulting in absences of four or more days in 2023 were included. The data were extracted from the Workplace Safety Site managed by the Ministry of Health, Labour and Welfare, Japan.^7^ This study adhered to the guidelines of the 1964 Declaration of Helsinki and was approved by the Ethics Committee of the University of Occupational and Environmental Health, Japan (No. R4-054).

The dataset included cases that were confirmed by local Labour Standards Inspection Offices based on legally mandated, employer-submitted reports.^8^ Occupational falls recorded in the dataset occurred between January 1 and December 31, 2023, and were finalized by the following April.^7^ The dataset contains detailed information on causative agents, types of accidents, nature of injury or illness, age (in five-year intervals), sex, and other relevant variables.

### Definition of occupational falls and sites

Occupational falls were defined as cases classified as “falls” under the accident type category in the dataset. In 2023, occupational falls accounted for 25.8% (36,058/139,998) of all registered cases of occupational accidents and diseases.

Fall sites were classified as either indoor or outdoor based on the causative agent category in the dataset. Indoor sites were further subcategorized into walkways, work platforms/walking planks, indoor stairs, and other indoor sites. Among these, walkways and work platforms/walking planks were defined as indoor level surfaces. Other indoor sites included roofs, beams, purlins, girders, trusses, buildings, structures, shoring, scaffolding, openings, and other temporary installations.

### Statistical analysis

Descriptive analyses were conducted to estimate the proportion of occupational falls by site of occurrence. Stratified analyses by sex and age were also performed. All analyses were performed using Stata version 17.0 (StataCorp, College Station, TX).

## Results

Among all occupational falls resulting in absences of four or more days, 63.2% occurred at indoor sites (Figure 1). The most frequently reported site was indoor level surfaces, accounting for 49.0% of all occupational fall cases, while indoor stairs accounted for 5.9% of all occupational fall cases (Figure 1).

**Figure 1.**
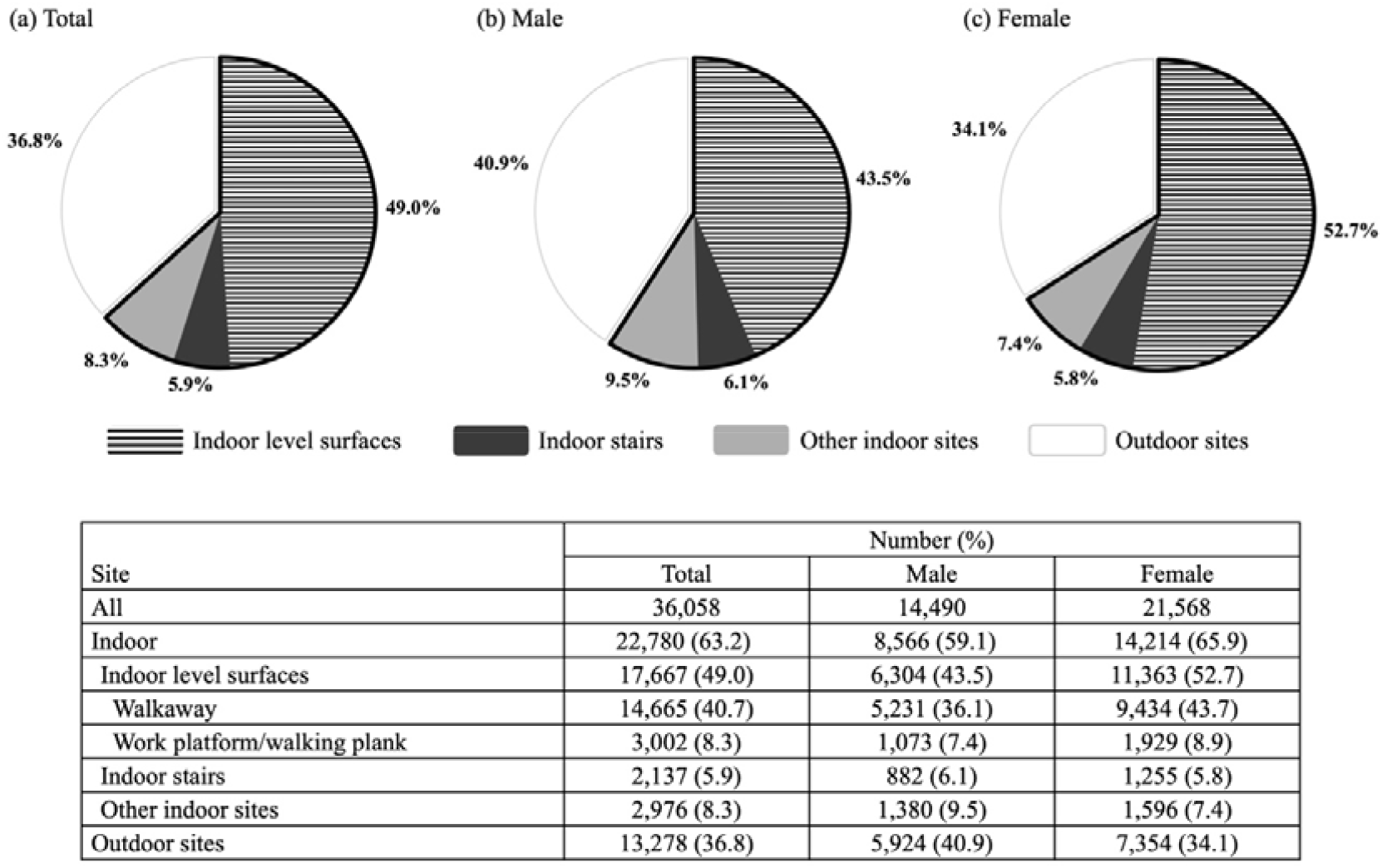
Occupational falls resulting in absences of four or more days by site of occurrence in Japan, 2023. The pie charts illustrate the proportions of occupational falls by site of occurrence within the workplace. Four categories are depicted: indoor level surfaces (horizontal stripes), indoor stairs (black-filled), other indoor sites (gray-filled), and outdoor sites (white area). The area enclosed by a bold black line represents the total proportion of indoor sites.

Stratified analyses by sex showed that the proportion of occupational falls at indoor sites was 59.1% among male workers and 65.9% among female workers (Figure 1). Falls on indoor level surfaces accounted for 43.5% in males and 52.7% in females. Similar distributions were observed across age groups (Table 1).

**Table 1.**
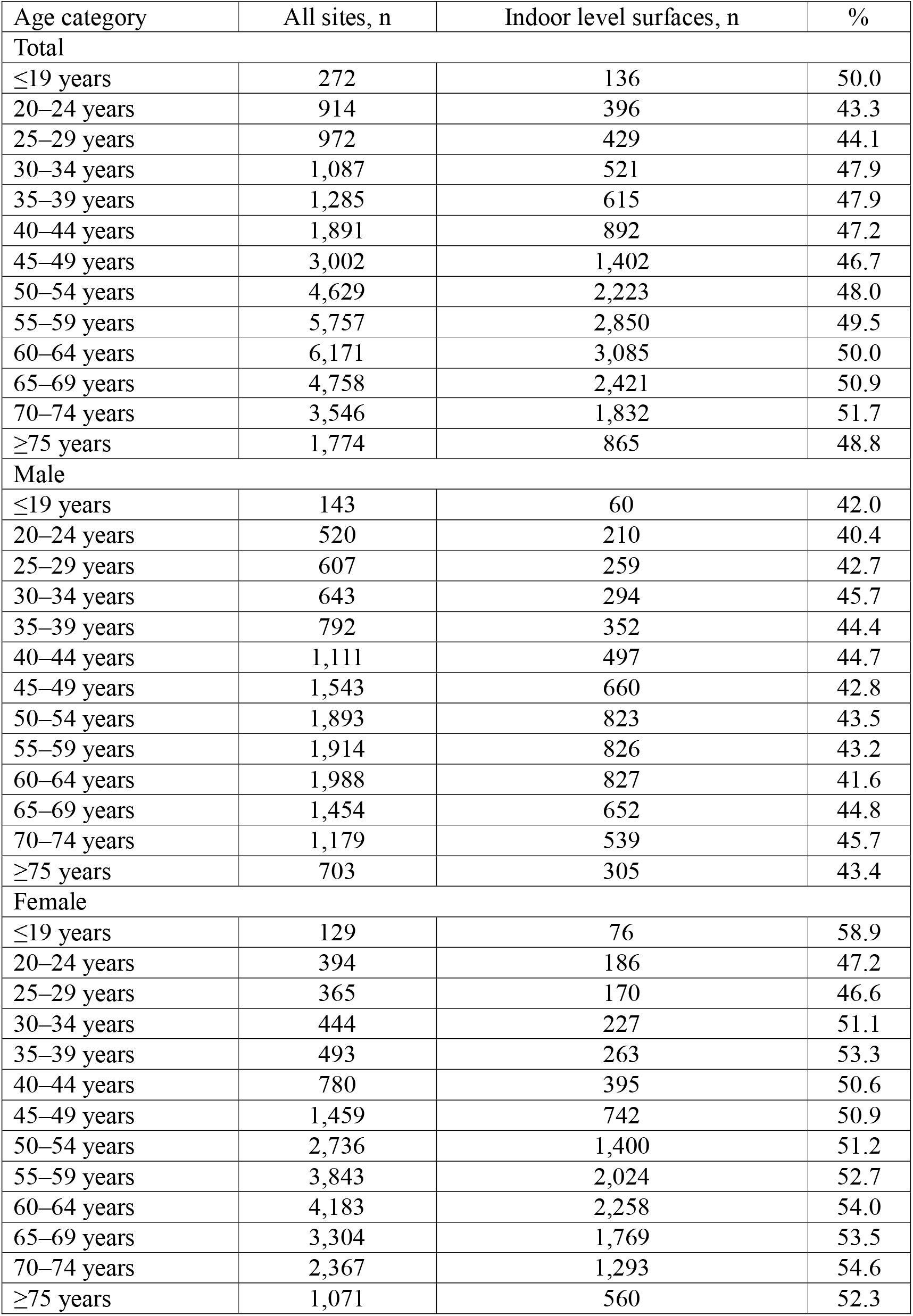
Occupational falls on indoor level surfaces by age group in Japan, 2023.

## Discussion

In Japan, we found that more than half of occupational falls occurred at indoor sites. Notably, indoor level surfaces accounted for approximately half of all cases, making them the most frequently reported site of occurrence. The proportion of occupational falls at indoor sites was 65.9% among female workers and 59.0% among male workers, with the difference largely attributable to a higher proportion of falls on indoor level surfaces among females. These findings indicate that occupational falls frequently occur in seemingly non-hazardous, controlled environments—particularly among female workers—and underscore the need for fall prevention strategies that extend beyond environmental modifications.

Although environmental modifications have traditionally been emphasized in preventing workplace falls, our findings suggest that this approach alone may be insufficient. National data revealed that many falls occurred on surfaces generally regarded as safe and controlled, such as indoor level surfaces. Therefore, complementary strategies targeting individual-level risk factors may be necessary.

For instance, improving physical functioning may help reduce fall risk. Static balance function—a proxy for occupational falls—tends to decline with age.^6^ Meanwhile, such deterioration may be mitigated through habitual physical activities, such as regular walking.^6^ Additionally, various lifestyle and behavioral factors, including smoking, sleep quality and patterns, hypertension, diabetes, and mental health, have also been associated with increased fall risk.^5,8,9^ Future research should further investigate the associations between theses modifiable risk factors and occupational falls.

This study has several limitations. First, it was descriptive in nature; thus, causal inferences regarding the mechanisms of occupational falls cannot be drawn. Second, although we analyzed the proportion of falls occurring on indoor level surfaces, the dataset lacked detailed contextual information on environmental risk factors, such as floor conditions or footwear at the time of the accident. Moreover, it was not possible to identify repeated fallers due to anonymization.

Despite these limitations, this study is the first to describe the distribution of occupational falls by workplace locations using a comprehensive national dataset. The use of data based on legally mandated employer-submitted reports under Japan’s Industrial Safety and Health Act ensures a high degree of completeness, accuracy, and generalizability. Furthermore, employers are obligated to report all occupational accidents and diseases, the risk of reporting or selection bias is likely minimal.

In conclusion, a substantial proportion of occupational falls in Japan occurred at indoor sites, particularly on indoor level surfaces. Given the limited potential for further conventional environmental interventions in such safe and controlled environments, future research and preventive strategies should focus more closely on individual-level risk factors to help mitigate the burden of occupational falls.^10^

## Acknowledgements

None.

## Data availability statement

The data are publicly available and can be accessed from the following URL: https://anzeninfo.mhlw.go.jp/user/anzen/tok/anst00.html.

## Author contribution statement

Conceptualization: Watanabe K, Hirohashi S, and Zaitsu M.

Funding acquisition: Zaitsu M.

Investigation: Watanabe K, Hirohashi S, Yoshimi T, and Zaitsu M.

Supervision: Zaitsu M.

Visualization: Watanabe K and Zaitsu M.

Writing-original draft: Watanabe K.

Writing-review & editing: Watanabe K, Hirohashi S, Yoshimi T, and Zaitsu M.

## Ethics approval and consent to participate

The study was approved by the Ethics Committee of the University of Occupational and Environmental Health, Japan (no. R4-054). The data were obtained from a publicly available government database and did not include any personally identifiable information. Therefore, informed consent and additional ethical procedures were not required.

